# The optimal level of serum vitamin D in apparently healthy Bangladeshi adult volunteer

**DOI:** 10.1101/2020.07.27.20162800

**Authors:** Anil Yadav, Shahjada Selim, Tahniyah Haq, Anil Kumar Shah, Md. Shahed Morshed, Md. Habibul Ghani, Ibrahim Faisal, Murshed Ahamed khan, Marufa Mustari, Mostafa Hasan Rajiv, M A Hasanat, Farid Uddin

## Abstract

**Introduction:** Vitamin D level has profound clinical implications but there is dilemma of optimal vitamin D cut off level among Bangladeshi population as well in many parts of the world. This study aimed to determine the optimal level of vitamin D in relation to intact parathyroid hormone (iPTH) and serum calcium in apparently healthy adult volunteer.

**Methods:** This observational cross-sectional study was carried out in 130 apparently healthy adult participants of BSMMU. All the subjects were taken their demographic profile and investigated for vitamin D level, iPTH, corrected calcium and phosphate. Serum 25(OH)D was measured by high performance liquid chromatography (HPLC) whereas iPTH, corrected serum calcium and serum phosphate were measured by chemiluminescent method.

**Results:** The mean 25(OH)D level was found to be 16.78(SD8.47) ng/ml and extensively distinct by age distribution and adequacy of sun exposure. There was substantially inverse correlation between serum iPTH and serum 25(OH)D (r = - 0.22, p = 0.01). Serum 25(OH)D levels < 27.5ng/ml were associated with a steep increase in serum iPTH levels. Serum iPTH was stabilized at level 54.5pg/ml by using the quadratic fit with plateau model.

**Conclusions:** From this study the optimal level of 25(OH)D for apparently healthy adult in Bangladesh is 27.5 ng/ml.

## Introduction

Vitamin D deficiency is currently recognized as a worldwide epidemic and its potential health implications are currently the subject of significant interest and controversy.^1^ There is no absolute consensus as to what a normal range for 25(OH)D should be. Many studies used inverse relationship between serum 25(OH)D and iPTH levels to determine normal serum 25(OH)D level. With the maximum efficiency of intestinal calcium absorption and adequate bone mineral density, iPTH concentrations began to plateau at their nadir.^2,3^ Different guidelines has set different cut off of optimal vitamin D status such as the Endocrine Society and many experts recommend > 30 ng/ml ^2,4^ and Institute of Medicine > 20 ng/ml.^5^ Bangladeshi patients are being treated following the guidelines of other population which may or may not be appropriate for our population. Thus, this study assesses the optimal range of serum 25(OH)D by testing the dynamic relationship between 25(OH)D and iPTH.

## Methods

### Study subjects

This Observational cross-sectional study was conducted in Department of Endocrinology, Bangabandhu Sheikh Mujib Medical University (BSMMU), Dhaka. Adult attendants of patients of BSMMU were requested to participate in the study. Participants who were taking or had received vitamin D or calcium supplements within last 120 days of sample collection, or took medication that affected calcium, vitamin D metabolism, and bone, or who had any known disease were excluded from study. The willing participants underwent full clinical assessment, prior to enrollment it was established that all of the participants had normal liver and kidney function. A total 130 apparently healthy adult attendants of patients of BSMMU were recruited during period of January 2018 to July 2019.

Anthropometric measurements were performed using same standardized techniques and calibrated digital scale (Seca, Germany), throughout the study. Body mass index (BMI) was calculated, for all subjects wearing light clothing and no shoes, as weight in kilograms (kg) divided by squared height in meters (m). Dietary intake of calcium and vitamin D was collected from 7-day food diaries. Physical activity information was obtained by International physical activity questionnaire (Short last 7 days self-administered format.^6^ Sunlight exposure was evaluated based on sunscreen use and daily average time of sun exposure between 1100 to 1500 hour with the sun exposure >20 % body surface.^7^

A morning fasting 10 ml venous blood sample was collected from each subject. Complete centrifugation of blood sample was done and serum was separated. Serum was collected in tubes and preserved at – 20°C until assay. Serum sample for calcium, albumin, phosphate and iPTH was transported to Department of Biochemistry, BSMMU where as serum sample for 25(OH)D was transported to Centre for Advanced Research in Sciences, Dhaka University. All the samples were analyzed by same method throughout the study period, using kits provided by the same manufacturer.

25(OH)D was measured by High performance liquid chromatography (SIL 20 series prominence HPLC, Shimadzu, Japan). The manufacturer’s normal range as reported in the kit was 20 to 100 ng/ml with analytical sensitivity 2.0 pg/ml. The coefficient of variability was 2.6-4.9%. iPTH was measured by solid phase two site chemiluminescent enzyme labeled immunometric assay, Immulite 2000 systems (Siemens, USA), automated analyzer (Beckman Coulter-AU680, Architect Plus ci8200). The manufacturer’s normal range as reported in kit was 12 to 65 pg/ml (1.3 to 6.8 pmol/L) with analytical sensitivity 3.0 pg/ml. The intra- and interassay precision (pg/ml) were 4.2 to 5.7% and 6.3 to 8.8% respectively. Serum calcium, phosphate, albumin, creatinine and alanine transaminase (ALT) were measured by chemiluminescent enzyme lableled immunometric assay using automated analyzer (Beckman Coulter-AU680, Architect Plus ci8200, Siemens, USA).

Serum calcium levels were adjusted for serum albumin using formula: Corrected serum calcium (mg/dl) = Serum Calcium + 0.8 * (40 - serum albumin mg/dl).^8^ Estimated glomerular filtration rate (eGFR) was calculated by CKD-EPI equations.^9^

Data was cross checked daily at the end of the day to find out errors and inconsistency of data and necessary editing, cleaning, coding and tabulation was done manually. All data were processed by SPSS program (version 22.0). Data expressed in frequencies or percentages for qualitative values and mean (±SD) for quantitative values. To compare the mean value of subgroups, independent t test, one way ANOVA were used as appropriate. Pearson’s correlation test was used to correlate between vitamin D, iPTH and other variables. The association between 25(OH)D and iPTH concentrations was studied both by linear and non-linear regression models. A quadratic model with plateau was fitted to model to see relationship between serum iPTH and serum 25(OH)D levels to objectively identify 25(OH)D level where iPTH reaches a plateau. p value ≤ 0.05 was considered statistically significant.

The study protocol was approved through the Institutional Review Board of BSMMU. A written informed consent was obtained from each participant.

## Result

The mean (SD) age of the participants was 37.57(12.22) years, ranging from 18 to 78 years.

Majority of participants (45.4%) were between 30 – 39 years while mean 25(OH)D level among various age distribution was found to be higher in 60 years and above 24.31±9.82. There were almost equal percentage of male (51.5%) and female (48.5%) included in the study participant with slightly higher mean 25(OH)D in female. The participants who reside on urban (74.6%) and rural (25.4%) area have mean value of 25(OH)D 16.33±8.44 and 18.08±8.55 respectively. Among occupation group mean value of 25(OH)D was low in student 13.63±7.90 and high among service holder group 18.72±8.17. The mean daily average time of sun exposure between 1100 hours to 1500 hours was 70±88.07 minutes but majority had inadequate sun light exposure. Those who had adequate sun exposure (18.5%) have mean 25(OH)D level 20.32±5.97 whereas inadequate sun exposure (81.5%) had 15.78±8.47. (Table 1)

**Table-1:**
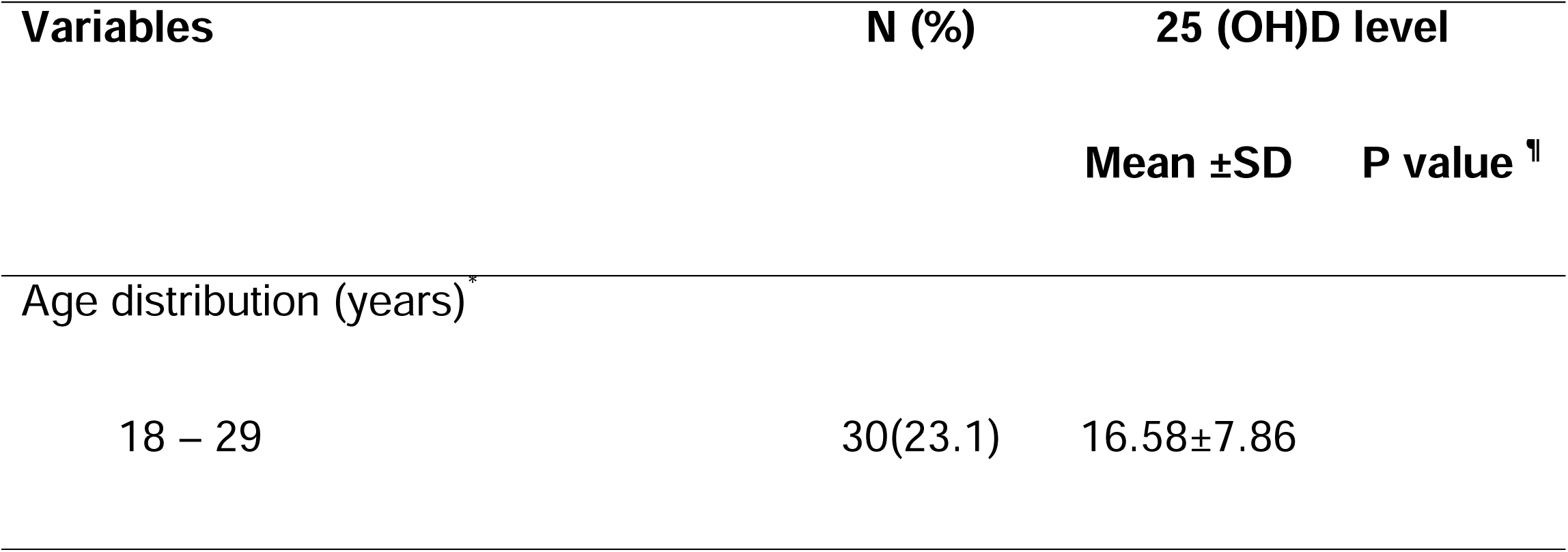
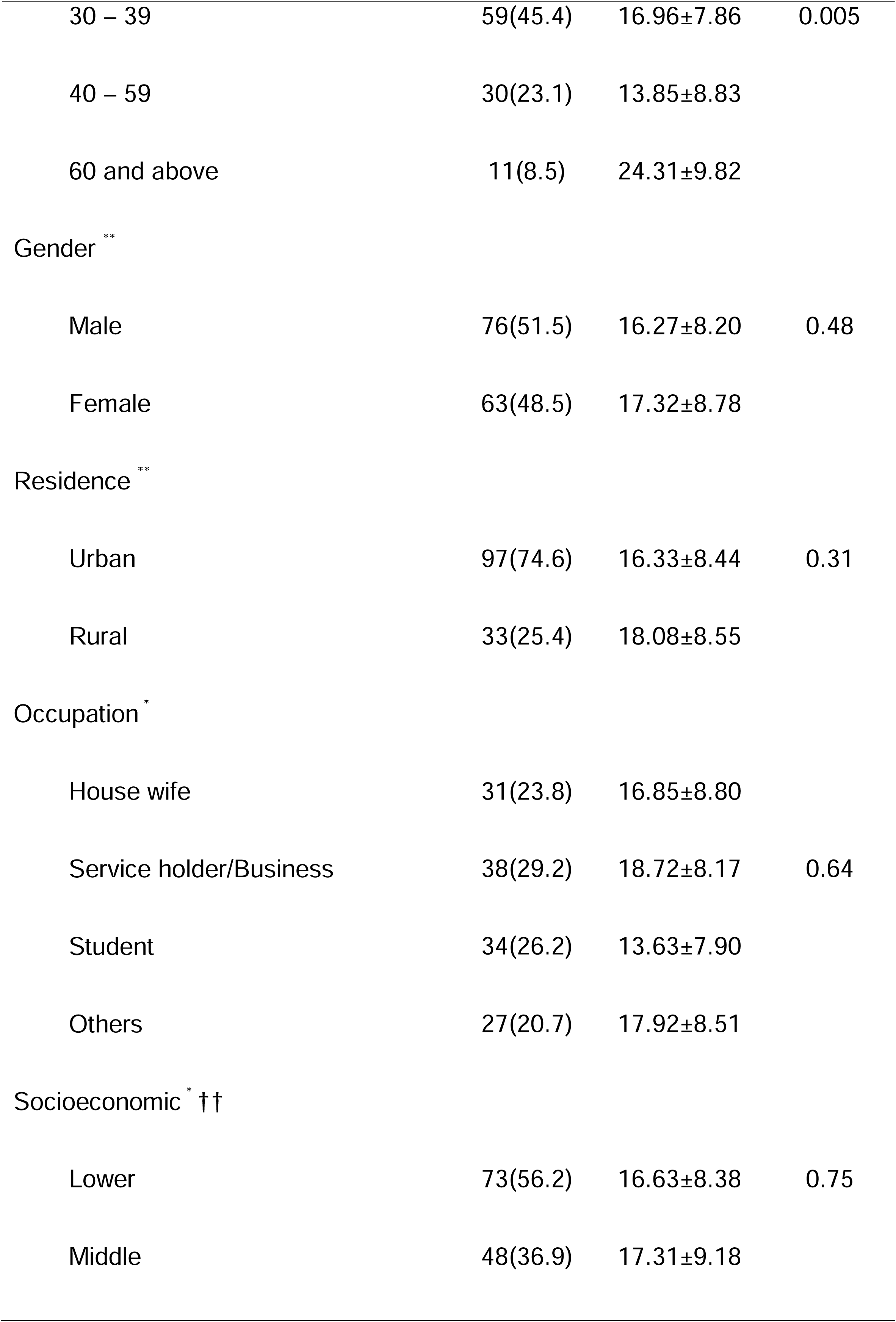
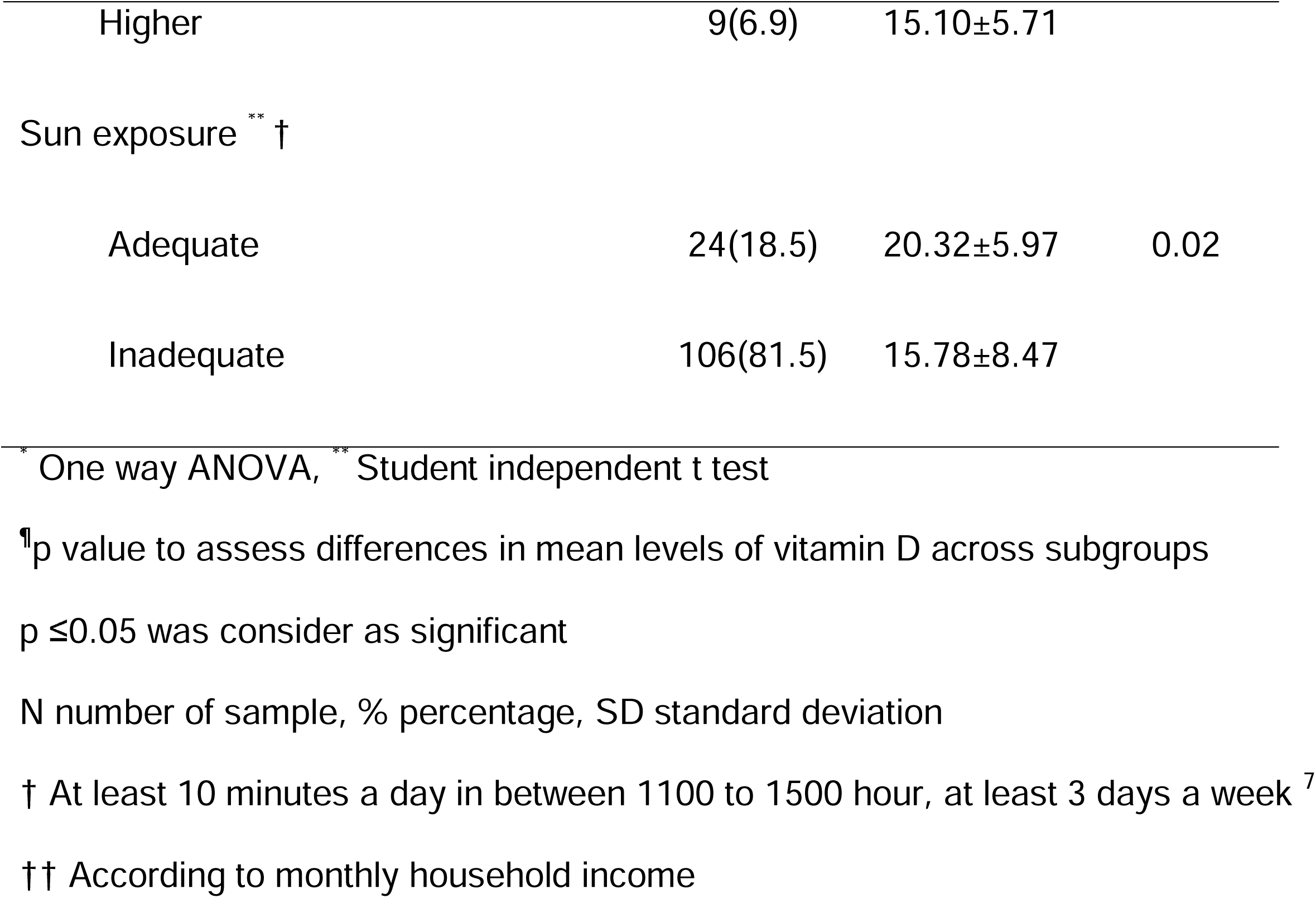
Socio demographic distribution of serum vitamin D among the study population (N=130)

| Variables | N (%) | 25 (OH)D level |  |
| --- | --- | --- | --- |
|  |  | Mean ±SD | P value <sup>†</sup> |
| Age distribution (years)<sup>*</sup> |  |  |  |
| 18 – 29 | 30(23.1) | 16.58±7.86 |  |
| 30 – 39 | 59(45.4) | 16.96±7.86 | 0.005 |
| 40 – 59 | 30(23.1) | 13.85±8.83 |  |
| 60 and above | 11(8.5) | 24.31±9.82 |  |
| Gender ** |  |  |  |
| Male | 76(51.5) | 16.27±8.20 | 0.48 |
| Female | 63(48.5) | 17.32±8.78 |  |
| Residence ** |  |  |  |
| Urban | 97(74.6) | 16.33±8.44 | 0.31 |
| Rural | 33(25.4) | 18.08±8.55 |  |
| Occupation * |  |  |  |
| House wife | 31(23.8) | 16.85±8.80 |  |
| Service holder/Business | 38(29.2) | 18.72±8.17 | 0.64 |
| Student | 34(26.2) | 13.63±7.90 |  |
| Others | 27(20.7) | 17.92±8.51 |  |
| Socioeconomic * †† |  |  |  |
| Lower | 73(56.2) | 16.63±8.38 | 0.75 |
| Middle | 48(36.9) | 17.31±9.18 |  |
| Higher | 9(6.9) | 15.10±5.71 |  |
| Sun exposure<sup>**</sup> † |  |  |  |
| Adequate | 24(18.5) | 20.32±5.97 | 0.02 |
| Inadequate | 106(81.5) | 15.78±8.47 |  |

\* One way ANOVA, \*\* Student independent t test
† p value to assess differences in mean levels of vitamin D across subgroups
p ≤ 0.05 was considered as significant
N number of sample, % percentage, SD standard deviation
† At least 10 minutes a day in between 1100 to 1500 hour, at least 3 days a week<sup>7</sup>
†† According to monthly household income

Overall, 63.8% of study participant reported moderate intensity physical activity; while mean 25(OH)D level was 18.86±8.47 among high intensity activity participants. All our study participants were consuming vitamin D enriched food like milk, eggs and fish in different proportion and amount. Almost half of the study participants had normal BMI while mean 25(OH)D was 18.40±7.68 among overweight (23.8%). (Table 2)

**Table-2:**
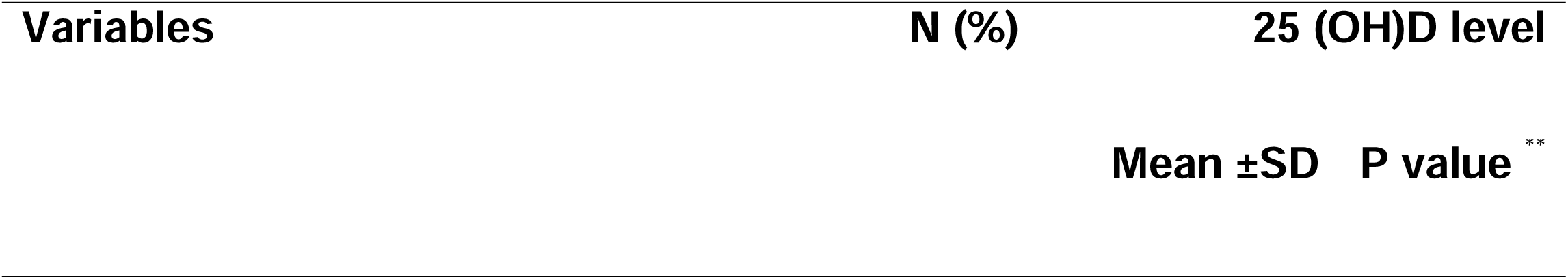
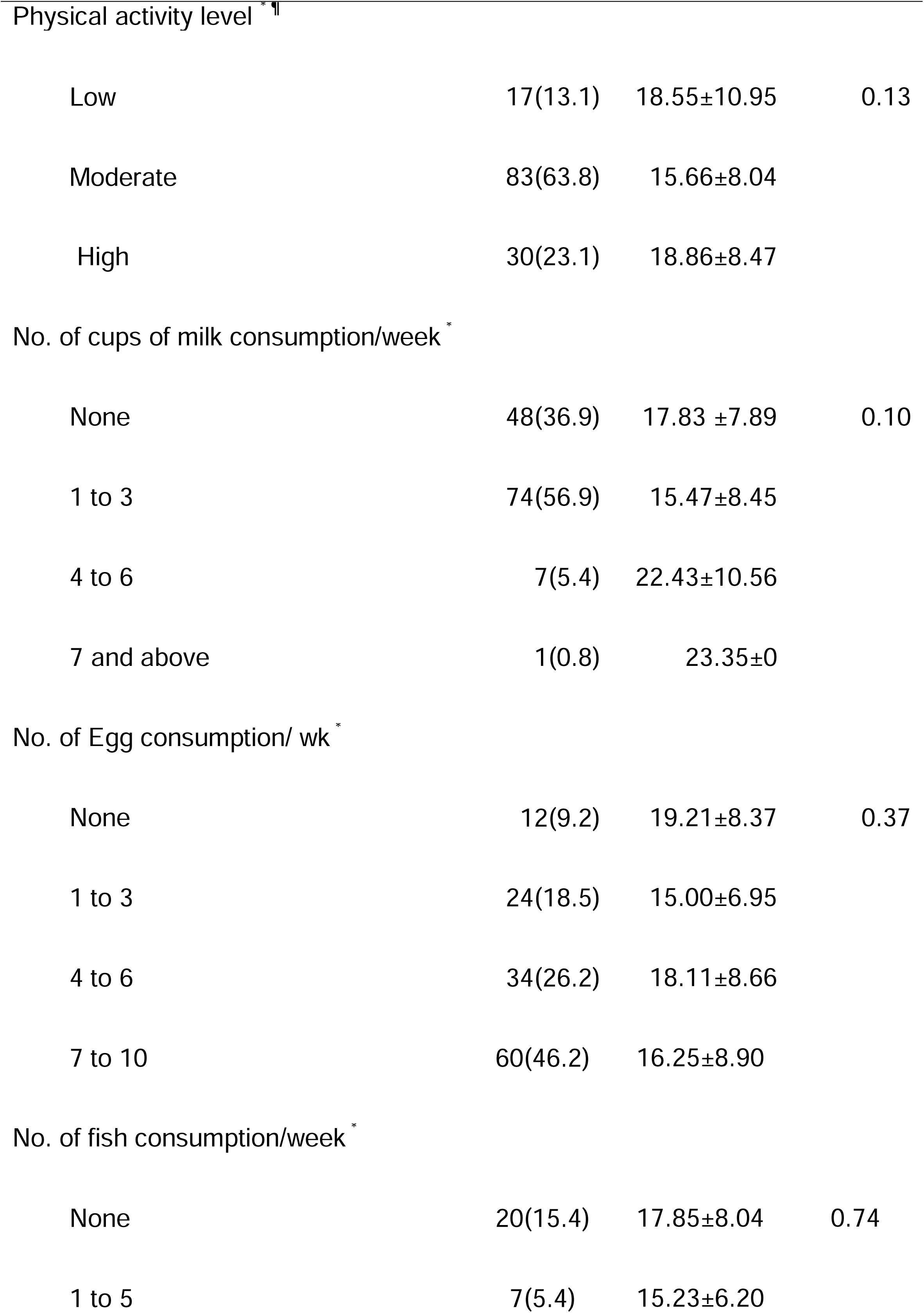
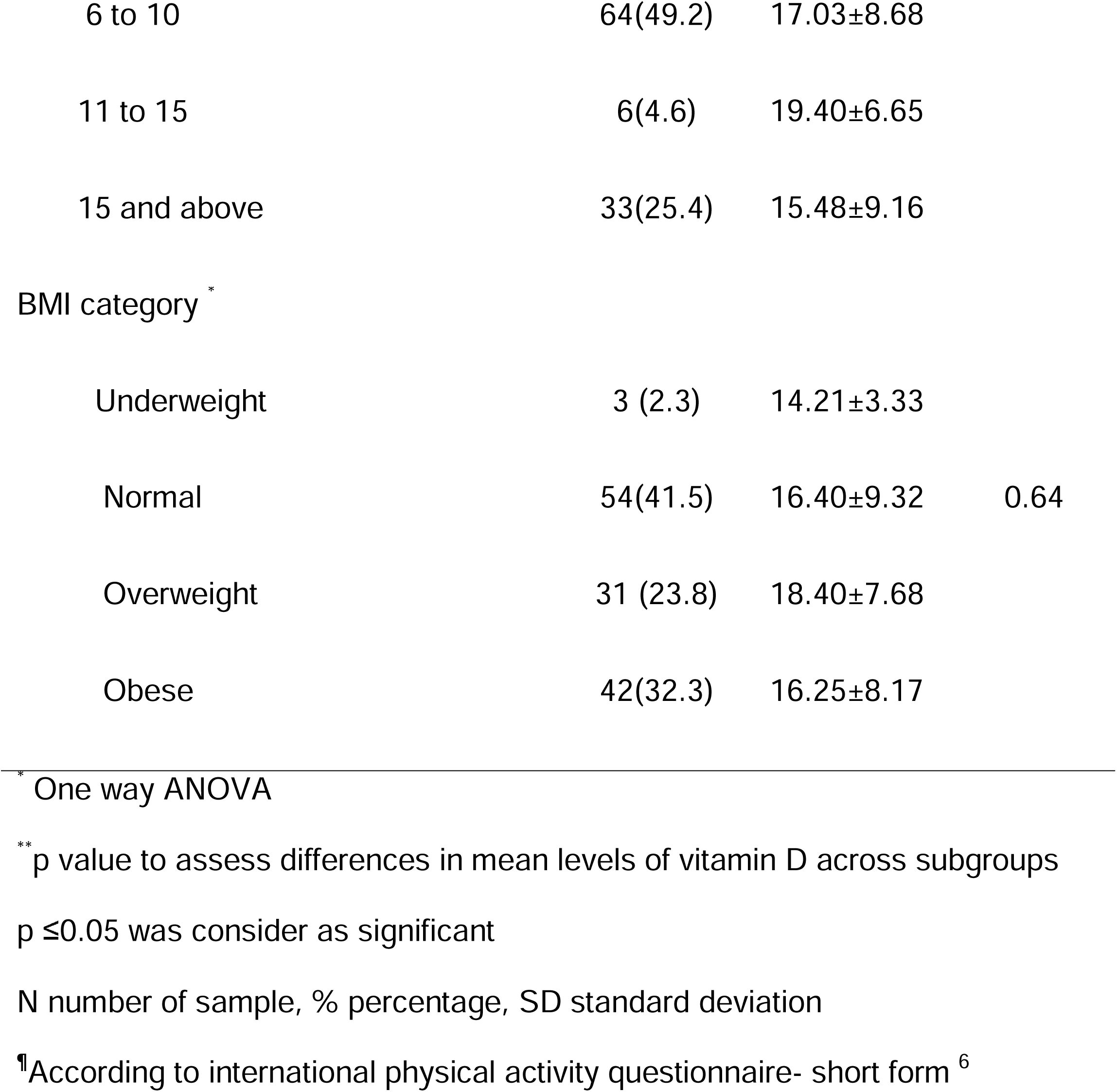
Distribution of serum vitamin D among the study population by level of activity, diet containing vitamin D and BMI (N=130)

The mean serum 25(OH)D, iPTH, corrected calcium and phosphate were 16.78±8.47, 60.68±23.01, 9.1±0.58 and 3.66±0.57 respectively.

None of the participants were below 5^th^ centiles whereas only 4.6% of the study participants were above 95^th^ centiles as shown in inter quartile group. Similarly, 24.61% of participants were between 0 to 25th, 25.38% between 25 to 50^th^ centiles, 25.38%, 50 to 75^th^ centiles and 24.61 % between 75 to 100^th^ centiles. Similarly figure 2 shows normal distribution of 25(OH)D in the study population.(Table 4)

**Table-3:**
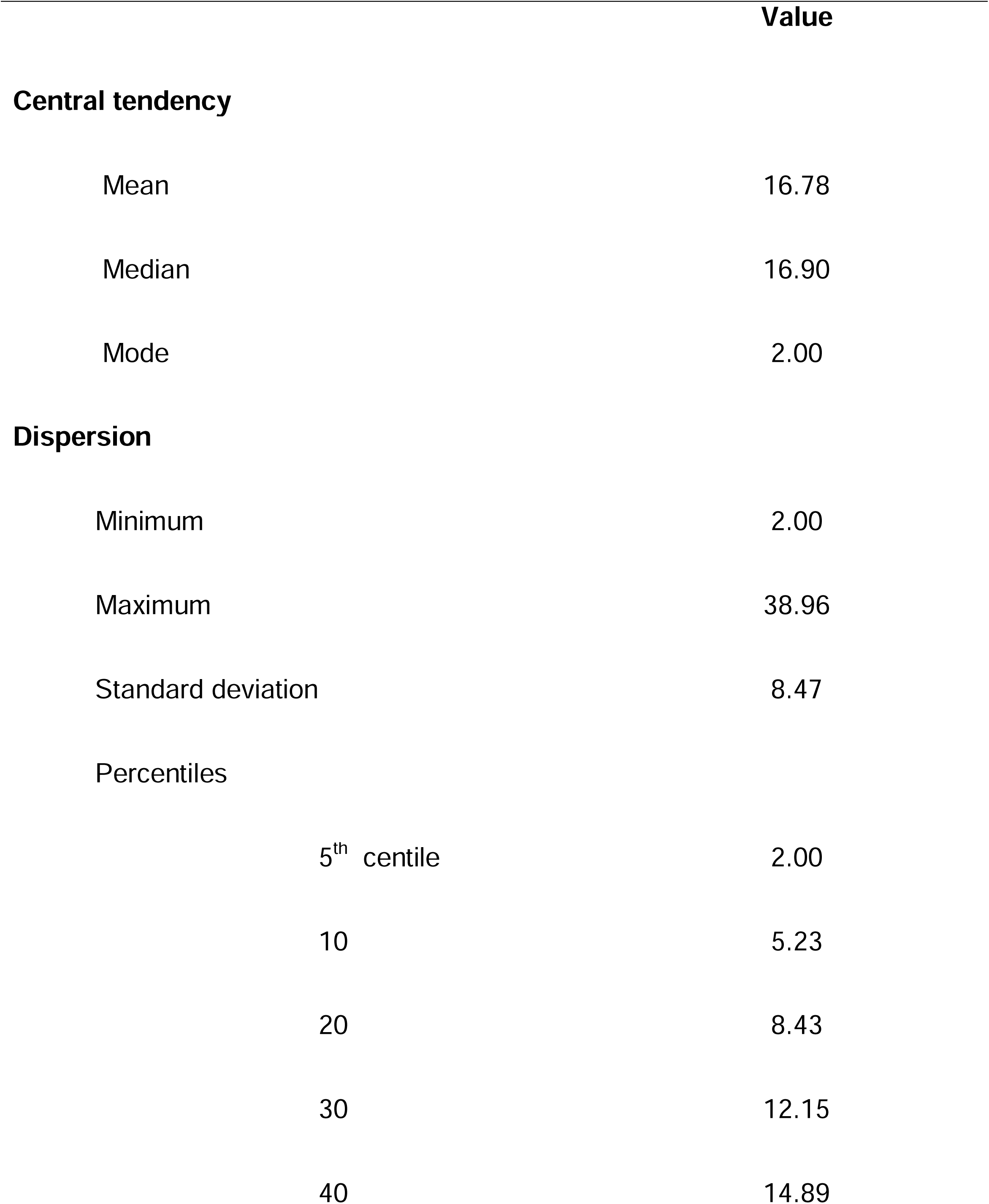
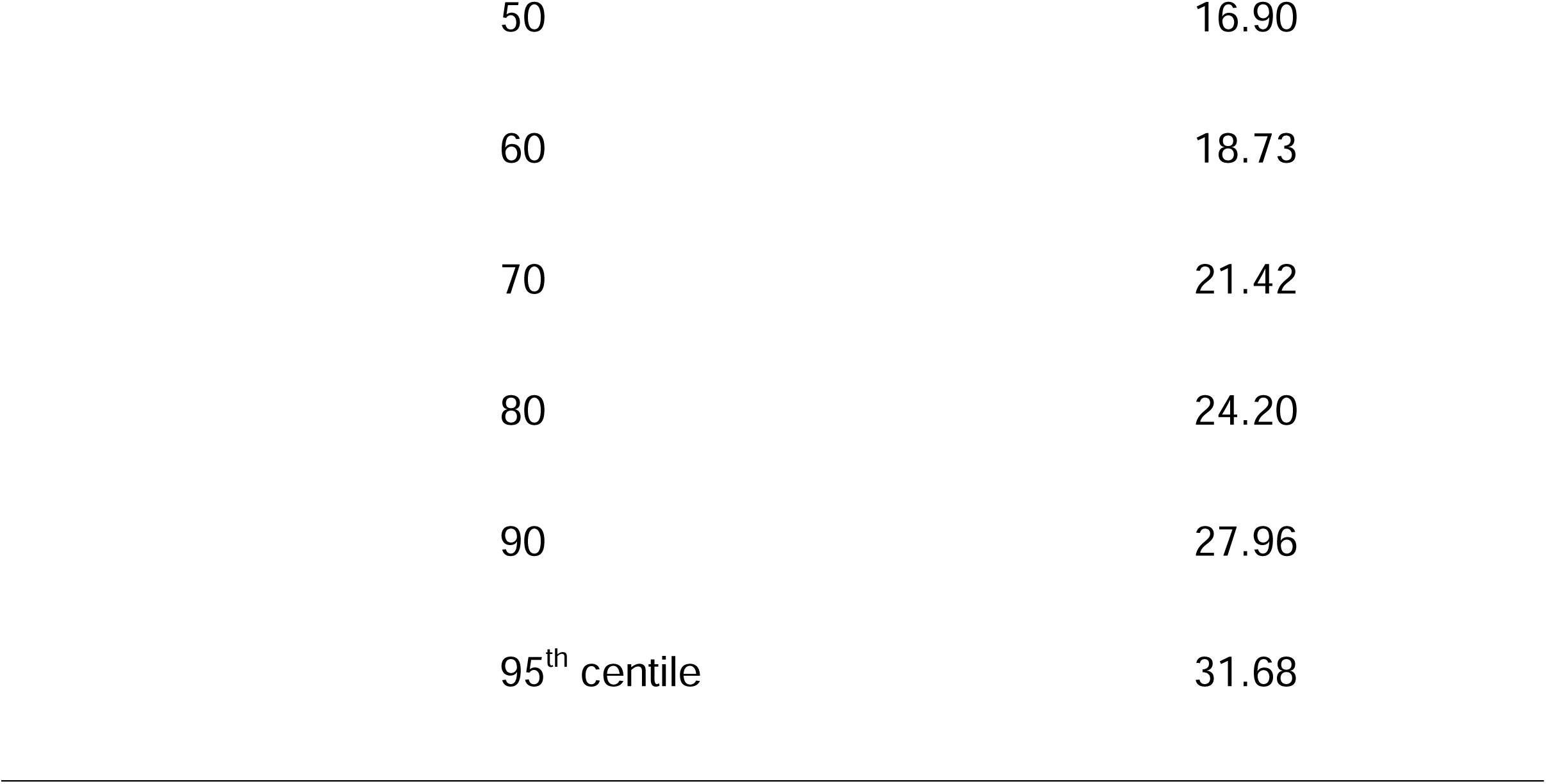
Statistics measures of central tendencies and variability of 25(OH)D of study population (N=130)

|  |  | Value |
| --- | --- | --- |
| **Central tendency** |  |  |
| Mean |  | 16.78 |
| Median |  | 16.90 |
| Mode |  | 2.00 |
| **Dispersion** |  |  |
| Minimum |  | 2.00 |
| Maximum |  | 38.96 |
| Standard deviation |  | 8.47 |
| Percentiles |  |  |
|  | 5<sup>th</sup> centile | 2.00 |
|  | 10 | 5.23 |
|  | 20 | 8.43 |
|  | 30 | 12.15 |
|  | 40 | 14.89 |

|  |  |
| --- | --- |
| 50 | 16.90 |
| 60 | 18.73 |
| 70 | 21.42 |
| 80 | 24.20 |
| 90 | 27.96 |
| 95<sup>th</sup> centile | 31.68 |

**Table-4:** Number of participants in inter quartile group (N=130)

| **Percentiles** | **N (%)** |
| --- | --- |
| 0 to 25 | 32(24.61) |
| 25 to 50 | 33(25.38) |
| 50 to 75 | 33(25.38) |
| 75 to 100 | 32(24.61) |
| Below 5<sup>th</sup> | 0 |
| Above 95<sup>th</sup> | 6(4.6) |

Serum 25(OH)D correlated positively with age, BMI and phosphate with correlation coefficient of 0.13, 0.23 and 0.002 respectively whereas negatively correlated with adequate sun exposure and iPTH with correlation coefficient of -0.20 and -0.22 respectively. Serum 25(OH)D was found to be significantly correlated with iPTH and adequate sun exposure. (Table 5)

**Table-5:**
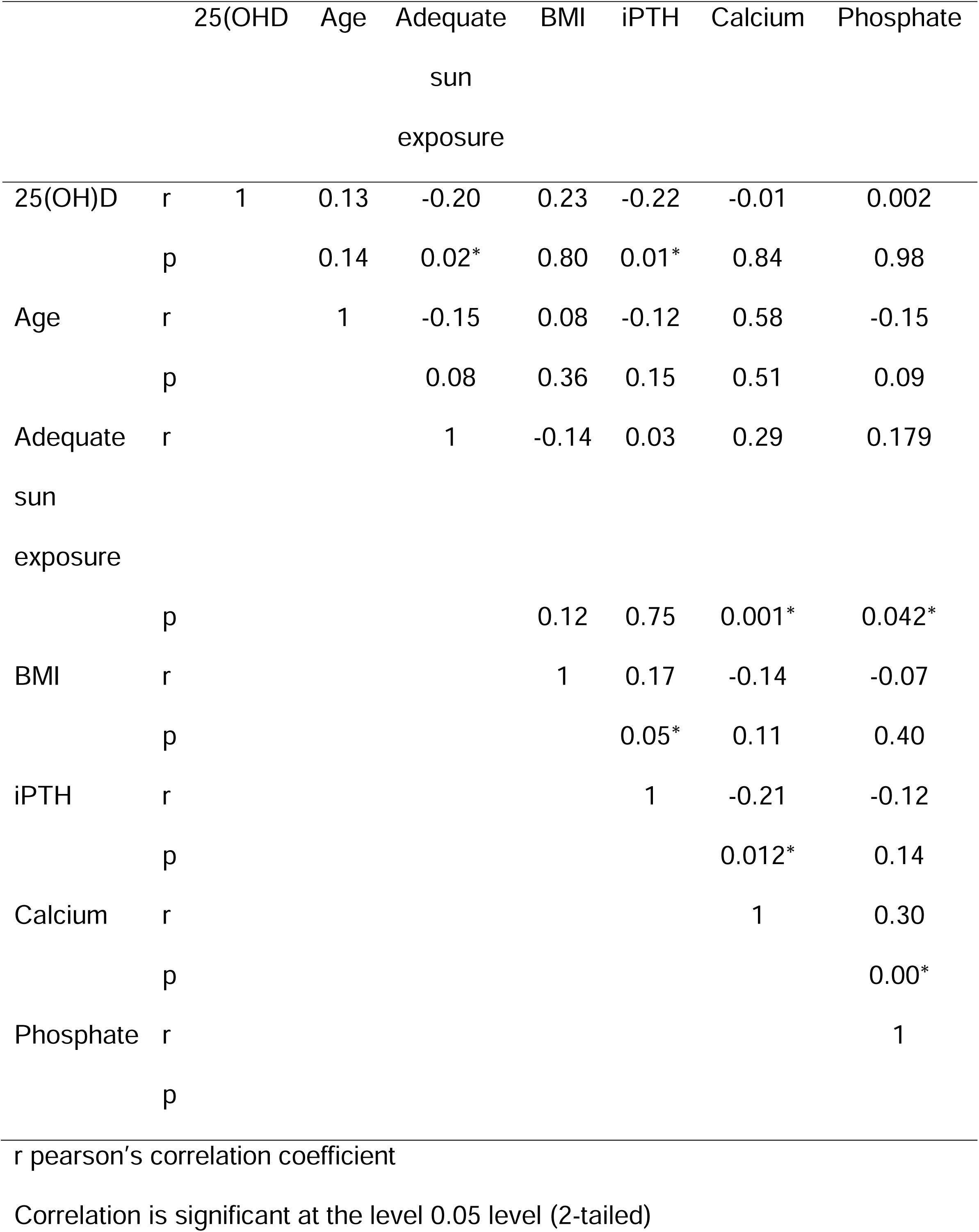
Pearson Correlations (r) of 25(OH)D with other investigated variables in study population (N=130)

|  |  | 25(OH)D | Age | Adequate<br>sun<br>exposure | BMI | iPTH | Calcium | Phosphate |
| --- | --- | --- | --- | --- | --- | --- | --- | --- |
| 25(OH)D | r | 1 | 0.13 | -0.20 | 0.23 | -0.22 | -0.01 | 0.002 |
|  | p |  | 0.14 | 0.02* | 0.80 | 0.01* | 0.84 | 0.98 |
| Age | r |  | 1 | -0.15 | 0.08 | -0.12 | 0.58 | -0.15 |
|  | p |  |  | 0.08 | 0.36 | 0.15 | 0.51 | 0.09 |
| Adequate<br>sun<br>exposure | r |  |  | 1 | -0.14 | 0.03 | 0.29 | 0.179 |
|  | p |  |  |  | 0.12 | 0.75 | 0.001* | 0.042* |
| BMI | r |  |  |  | 1 | 0.17 | -0.14 | -0.07 |
|  | p |  |  |  |  | 0.05* | 0.11 | 0.40 |
| iPTH | r |  |  |  |  | 1 | -0.21 | -0.12 |
|  | p |  |  |  |  |  | 0.012* | 0.14 |
| Calcium | r |  |  |  |  |  | 1 | 0.30 |
|  | p |  |  |  |  |  |  | 0.00* |
| Phosphate | r |  |  |  |  |  |  | 1 |
|  | p |  |  |  |  |  |  |  |

r pearson's correlation coefficient
Correlation is significant at the level 0.05 level (2-tailed)

In the linear regression analysis, one unit increase in the 25(OH)D concentration on the decreased iPTH levels by 0.22units (p= 0.02).(Table 6)

**Table-6:** Simple linear regression showing the predictive association of 25(OH)D with iPTH (N=130)

|  | **R<sup>2</sup>** | **B** | **Beta** | **Coefficient** | **p** | **95 % CI** |
| --- | --- | --- | --- | --- | --- | --- |
| **Vitamin D** | 0.048 | 70.46 | - 0.22 | -0.547 | 0.012 | 61.78 – 79.15 |

R<sup>2</sup> Coefficient of determinants
CI confidence interval
$p \leq 0.05$ was consider as significant

Based on the relationship between 25(OH)D and iPTH concentrations, the plateau for iPTH concentration was reached at 54.5 pg/ml. We observed that 25(OH)D concentrations higher than about 27.5 ng/ml were required to keep iPTH concentrations low. (Figure-4)

**Figure-1:**
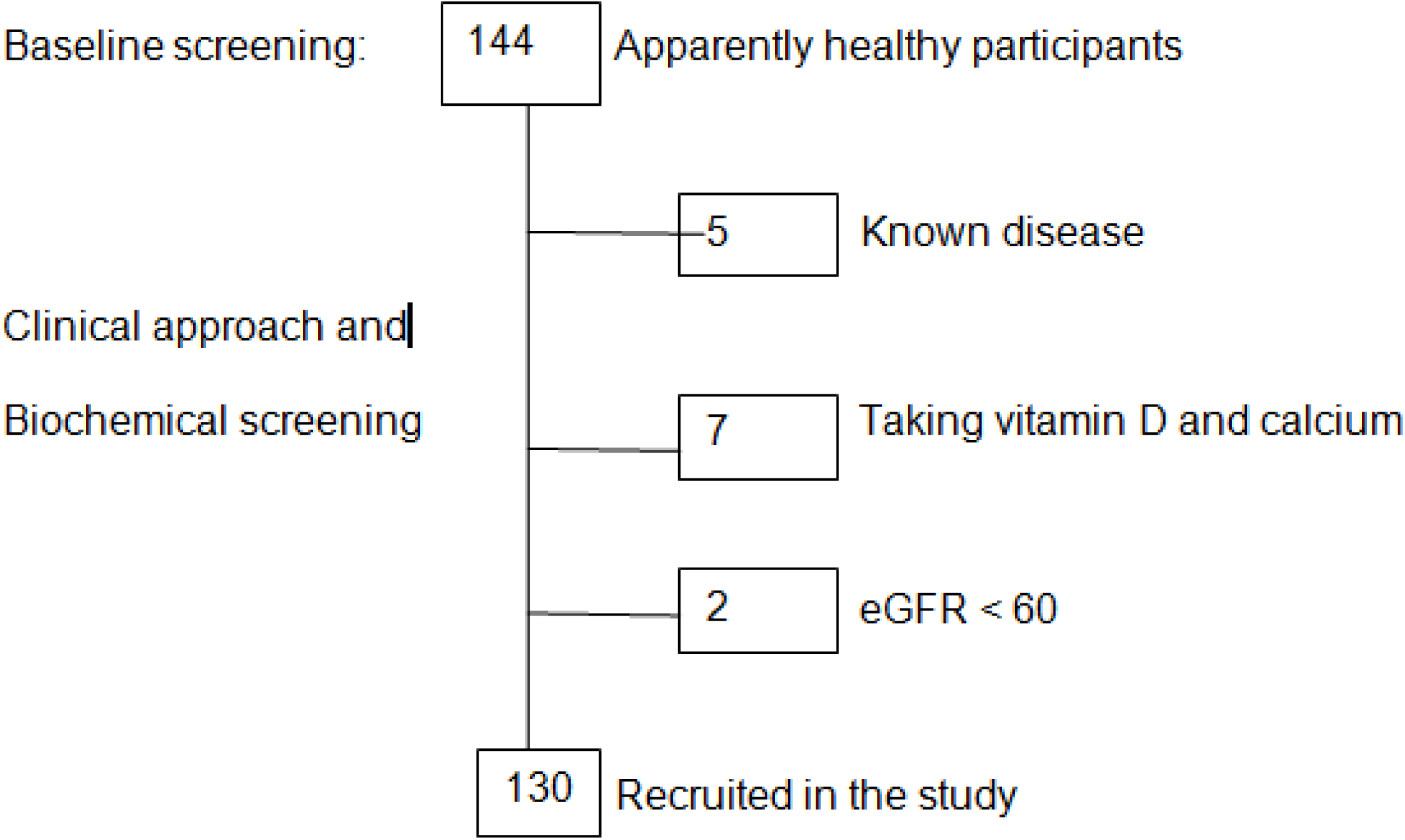
Patient recruitment.

**Figure-2:**
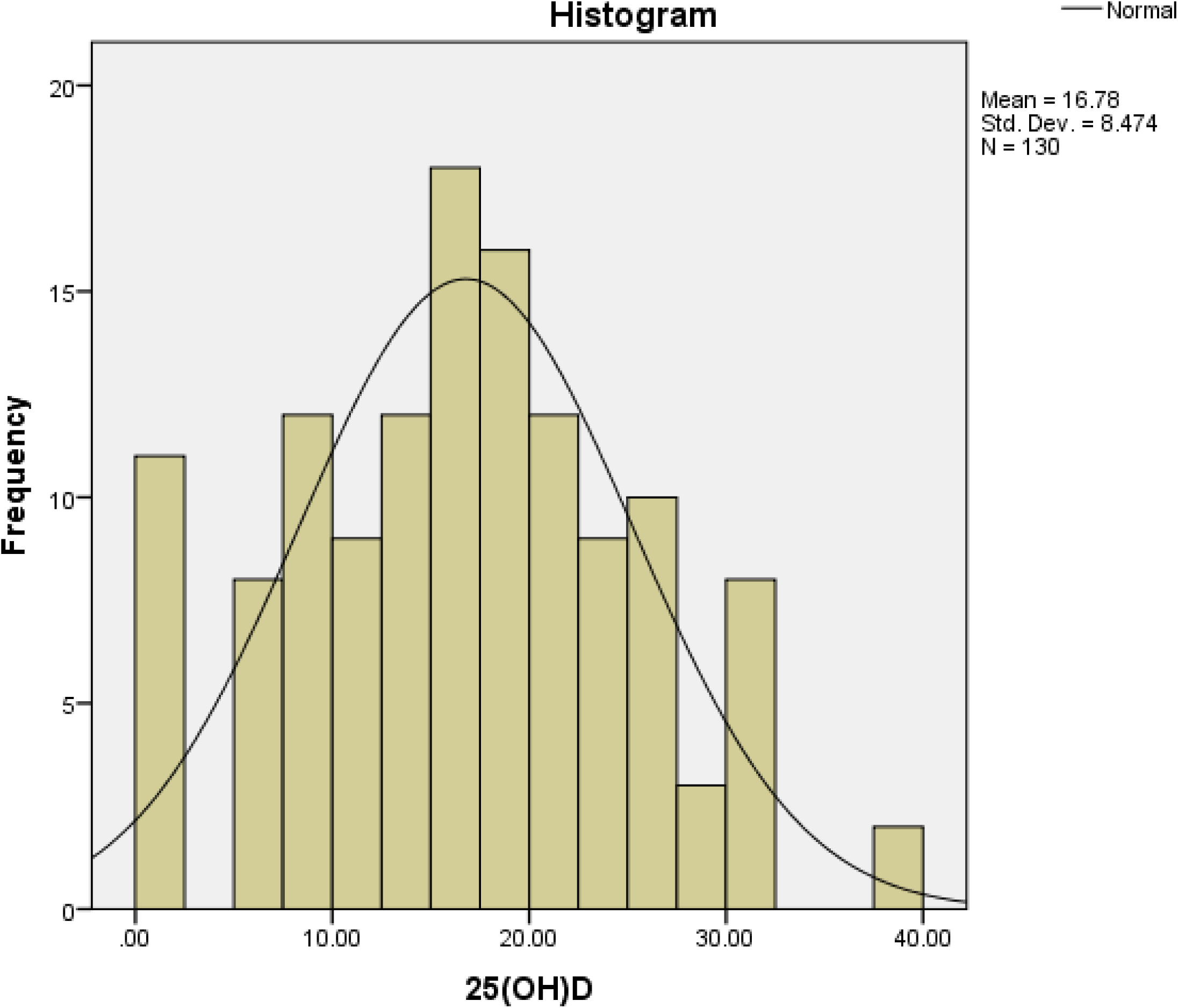
Distribution of 25(OH)D in the study population.

**Figure-3:**
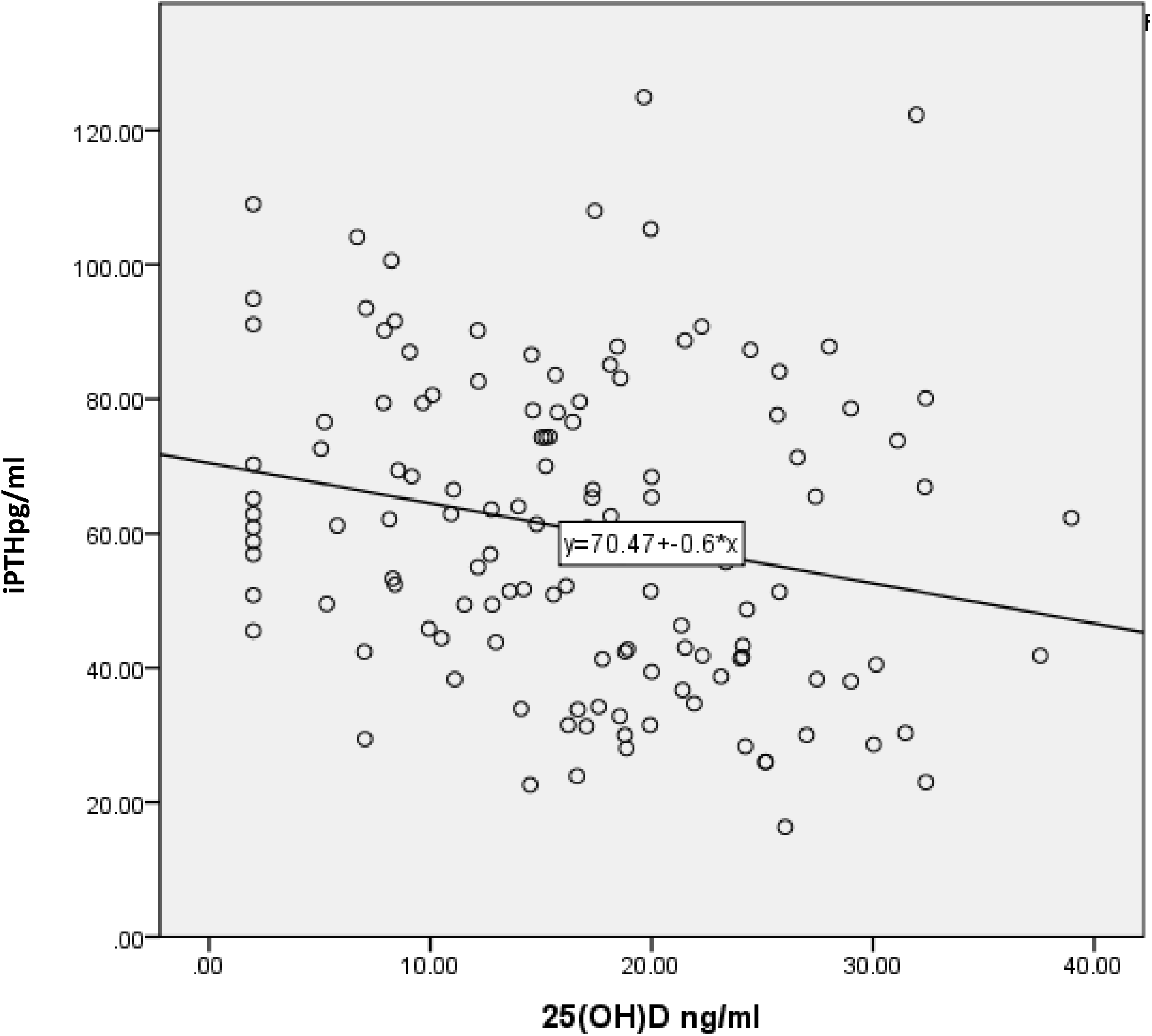
Correlation between 25(OH)D and iPTH in the study population.

**Figure-4:**
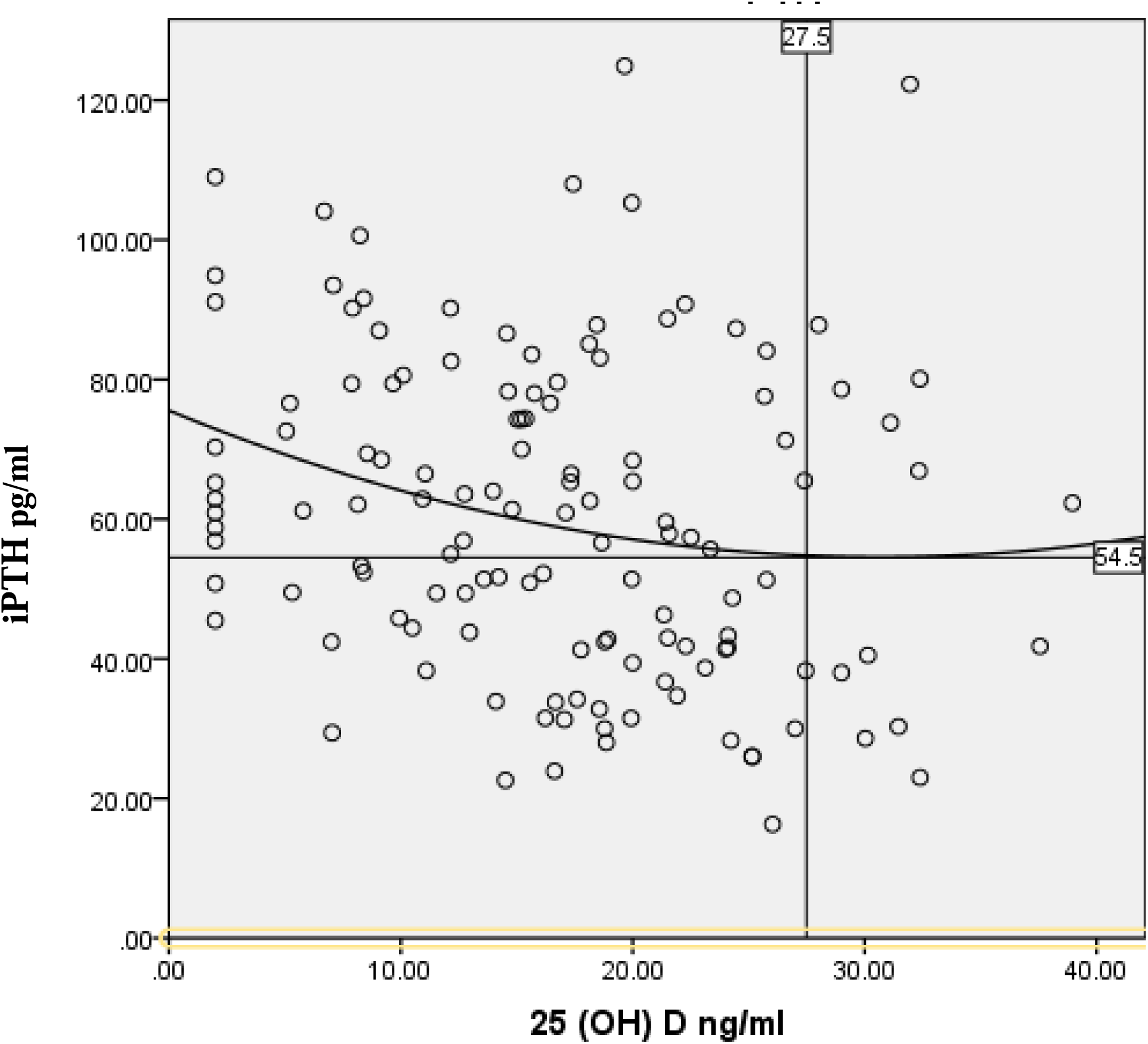
Association between serum 25(OH)D and iPTH in quadratic fit model.

The optimal 25(OH)D level was found to be 27.5 ng/ml in this study. Of the total study population, 90% had 25(OH)D below 27.5 ng/ml. According to the Endocrine society classification, 25(OH)D < 30 ng/ml was present in 92.3% and 66.2% of study population had 25(OH)D level below 20 ng/ml which was the cut off value as per IOM classification. (Figure 5).

**Figure-5:**
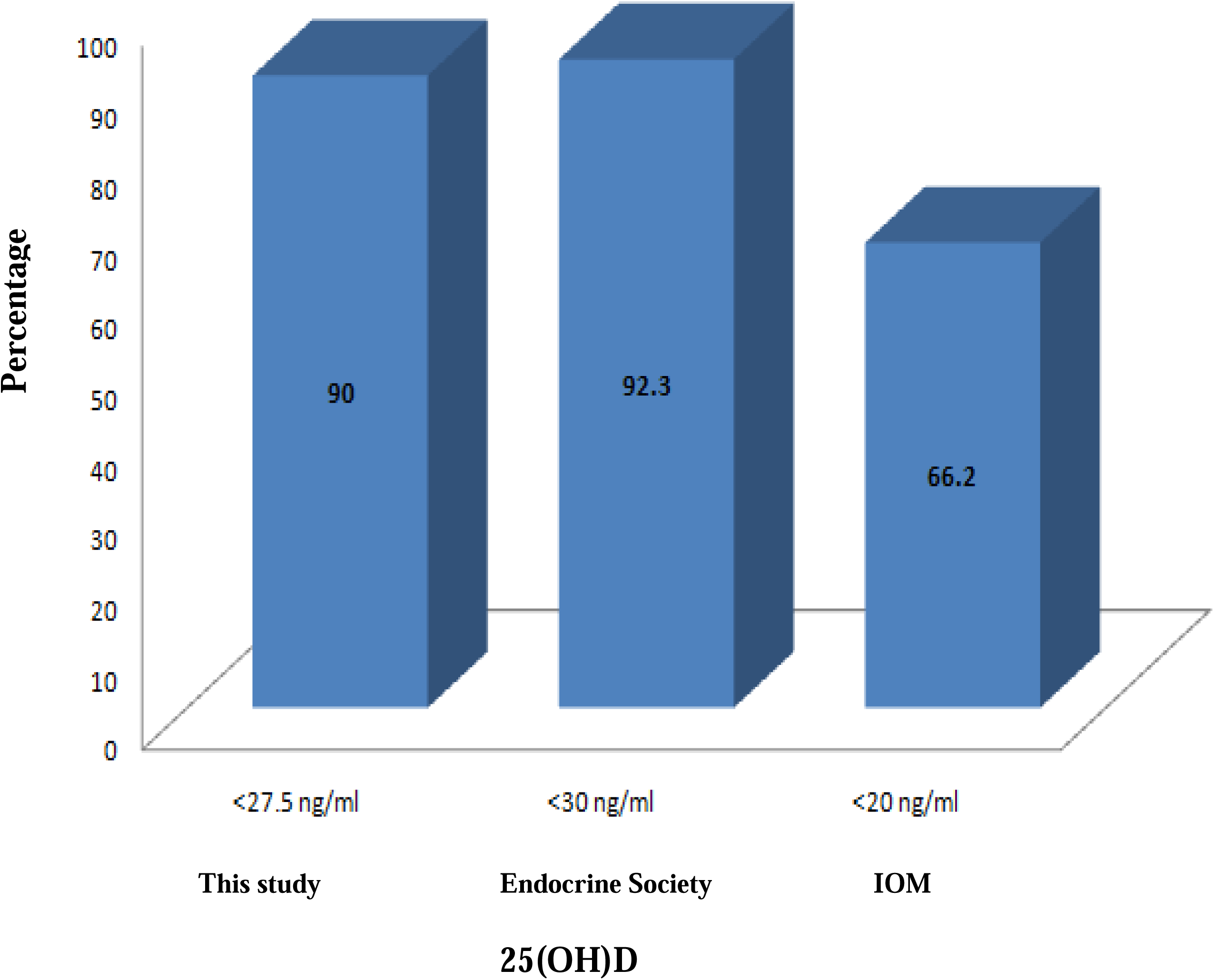
Distribution of the study population by vitamin D status in comparison to the Endocrine society and IOM classification (N=130).

## Discussion

This study showed that mean serum 25(OH)D was 16.78±8.47 with the optimal level approximately 27.5 ng/ml, measured by HPLC method, in apparently healthy study population with normal kidney function and normal serum calcium levels. This was based on the level at which serum iPTH plateaus and/or is maximally suppressed. Several studies from sunny countries reported a reference range of serum 25(OH)D 54 – 90 ng/ml to indicate vitamin D sufficiency in the normal subjects.^10^

In this study, 90% of study participant had serum 25(OH)D level below 27.5 ng/ml. According to US Endocrine Society we found 92.3% of population had low 25(OH)D with cut off < 30 ng/ml but according to IOM only 66.2 % of study population had low 25(OH)D with cut off < 20 ng/ml. We found a significant inverse relationship between serum 25(OH)D concentration and iPTH (r= -0.22 and p = 0.01) which was similar with study conducted on women of Jakarta and kuala Lumpur and half of Israeli population.^11,12^ Another study conducted on female garment worker of Bangladesh,^13^ observed similar significant inverse relationship between 25(OH)D and iPTH (r= - 0·25, p< 0·001). Based on the relationship between serum 25(OH)D and iPTH concentration, this study found that the plateau for the iPTH concentration by quadratic model was reached at 54.5 pg/ml. However, due to heterogeneous and small population the plateau level was not consistent. We observed that 25(OH)D concentrations higher than 27.5 ng/ml were required to keep iPTH concentrations within normal limit. According to the study conducted on female garment workers in Bangladesh,^13^ the plateau for iPTH concentration was reached at 21 ng/l when 25(OH)D reached 15.2 ng/ml, which was not consistent with our study. The reason may be decreased cutaneous vitamin D synthesis due to inadequate sun exposure. The optimal level of 25(OH)D (27.5 ng/ml) of the participant was almost similar to the Endocrine Society (> 30 ng/ml) based on elevated iPTH that was consistently lowered to a plateau when serum 25(OH)D was at 30ng/ml or higher. However, the IOM defined optimal level of 25(OH)D (>20ng/ml) based on human requirement of vitamin D for the general population in context of its relevance to bone accretion, bone maintenance and bone loss.^5^ Substantial studies found that 25(OH)D concentrations from about 12 to 50 ng/ml (30 to 125 nmol/l) were required to maintain a normal iPTH level.^10,14^ In the Israeli population, serum 25(OH)D levels <20 ng/ml were associated with a steep increase in iPTH levels, which lessen with increasing 25(OH)D levels and reached a plateau at 25(OH)D levels of 30 to 34 ng/ml.^12^ It was appraised that iPTH began to increase when serum 25(OH)D level was< 31.56 ng/ml. This point corresponded to a serum iPTH level of 62.5 pg/ml in all cases tested. In French population,^15^ iPTH levels began to plateau at their nadir when 25(OH)D levels were between 30 and 40 ng/ml. Healthy individuals in Australia and Riga, serum 25(OH)D of 38 ng/ml was sufficient to avoid rise in iPTH.^16,17^ Our neighbor country India had systematic review on vitamin D level in apparently healthy Indian population of 40 study had shown the overall effect size of serum 25(OH)D levels among Indians as 14.16 ng/ml (confidence interval [CI]: 13.27–15.05). However, they didn’t observe the iPTH level.^18^ Among young Lebanese people iPTH reached a plateau was maximally suppressed when 25(OH)D was 10 ng/ml or less.^19^

Vitamin D status also allies to age and gender^11,20^ A study in Canada found that vitamin D status among the 60−79 age group was higher comparing among the 20 – 30 youth age group.^21^ In our study, also the result showed that mean serum 25(OH)D concentrations in age 60 and above was 24.31±9.82 which was higher than 18 – 29 and 30 – 39 age group. However, this study revealed 40 – 59 age group had the lowest 13.85±8.83 25(OH)D level. A tenable explanation could be that most of them were working in the indoor setting. As a result, they spent less time outside, hence less sun exposure. In addition, they did not have the adequate habit of drinking milk, eating fish and meat.

Female gender are one of the most frequently reported risk factors for hypovitaminosis D but in contrast we found almost similar mean 25(OH)D level in both male and female 16.27±8.20 and 17.32±8.78 respectively, with level in males being slightly higher. ^22^ A study from the Netherlands by found that, serum 25(OH)D concentrations of males was higher than that of females which was consistent with our result. ^23^ The reason being, that the males did more outdoor activities and ate more varieties of foods than females. Likewise in Middle-Eastern young population they found mean 25(OH)D levels in overall population was 31±12.48 ng/ml with a lower mean value in men compared with women (29.01±11.23 ng/ml in men versus 33.2±13.4 in women), p=0.001.^24^

Exposure to the sunlight is essential to obtain adequate vitamin D, as it is mainly produced in skin by exposure to UVB radiation from the sunlight.^20^ Timing and proper skin exposures are also important for vitamin D photosynthesis. In this study, only 18.5 % gave history of proper sun exposure whereas 81.5% study population had inadequate sun exposure as most of them spent a major part of day time in office building as service holder and housewife 29.2% and 23.8% respectively. We presume and agree with the authors who indicated that low intensity of the sun in the morning, shaded sun shines, shadows of tall buildings and trees, upright position of the subjects, high pollution in the air, covered-up-dressing style as well as the dark skin necessitate the need for prolonged exposure for adequate synthesis of vitamin D in the skin.^25^

In this study, the intake of selected nutrition D containing food did not show significant correlation with Vitamin D. A similar finding was also found in investigation where authors suggested that about 10% of vitamin D is derived from dietary sources indicating that dietary intake of vitamin D is a relatively poor predictor of overall vitamin D status.^26,27^ Insignificant affiliation between dietary vitamin D intake and serum level of 25(OH)D has been shown in studies performed in Europe^28,29^ Bangladesh currently does not fortify any foods with vitamin D.^30^

Therefore, we would conclude that there is no universally accepted optimal range or cut-off value for 25(OH)D available to define vitamin D deficiency or insufficiency. Moreover, the demarcation line between vitamin D sufficiency and insufficiency is not clearly defined.^18^ The continuing deliberations have raised the concerns regarding normal cutoff value of vitamin D level as well as the need for vitamin D supplementation in Bangladeshi population. Further, studies are needed to establish the normal value for serum 25(OH)D level in Bangladeshi population.

There were a number of limitations in our study. The samples for all types of measurements could not be run in single assay to avoid interassay variation of coefficiency. We were unable to correlate with bone mineral density, biochemical bone markers and maximal efficiency of intestinal calcium absorption.

## Conclusion

The optimal level of serum 25(OH)D for apparently healthy adult in Bangladesh is 27.5 ng/ml. Study encompassing people from different groups and measuring serum vitamin D round the year, may yield more robust level of optimal serum vitamin D. Large scale study recruiting homogenous Bangladeshi adult population should be carried out

## Data Availability

The data that support the findings of this study are available from the corresponding authorupon reasonable request.

## Acknowledgement

Department of Biochemistry, Bangabandhu Sheikh Mujib Medical University, Dhaka and Center for Advance Research in Sciences, Dhaka University, Dhaka, Bangladesh for their support in biochemical analysis. Beximco Pharmaceuticals Limited, Bangladesh for technical support.

---

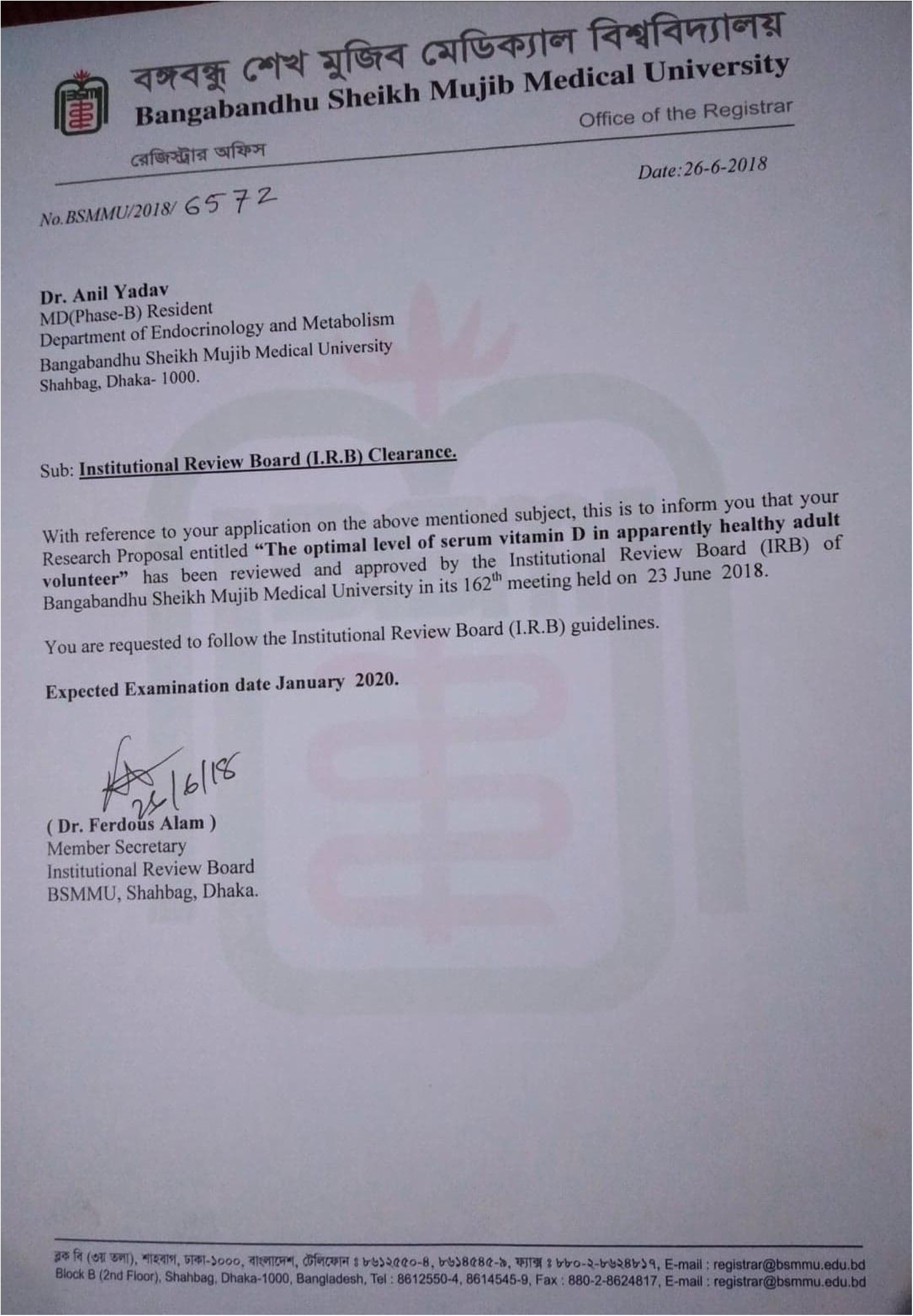

## Notes

### Competing Interest Statement

The authors have declared no competing interest.

### Author Declarations

Research entitled "The optimal level of serum vitamin D in apparently healthy Bangladeshi adult volunteer" was reviewed and approved by Institutional review board (IRB) of Bangabandhu Sheikh Mujib Medical University (BSMMU) in its 162 meeting held on 23 June 2018. Reference no. BSMMU/2018/6572

